# Elevated D-Dimer Levels are Associated with Increased Risk of Mortality in COVID-19: A Systematic Review and Meta-Analysis

**DOI:** 10.1101/2020.04.29.20085407

**Authors:** Siddharth Shah, Kuldeep Shah, Siddharth B Patel, Forum S Patel, Mohammed Osman, Poonam Velagapudi, Mohit K. Turagam, Dhanunjaya Lakkireddy, Jalaj Garg

**Author notes:** Corresponding Author, Jalaj Garg MD FACC FESC, Division of Cardiology, Cardiac Arrhythmia Service, Medical College of Wisconsin, 10000 Innovation Drive, Milwaukee, WI 53226.

## Abstract

**Introduction:** The 2019 novel Coronavirus (2019-nCoV), now declared a pandemic has an overall case fatality of 2–3% but it is as high as 50% in critically ill patients. D-dimer is an important prognostic tool, often elevated in patients with severe COVID-19 infection and in those who suffered death. In this systematic review, we aimed to investigate the prognostic role of D-dimer in COVID-19 infected patients.

**Methods:** We searched PubMed, Medline, Embase, Ovid, and Cochrane for studies reporting admission D-dimer levels in COVID-19 patients and its effect on mortality.

**Results:** 18 studies (16 retrospective and 2 prospective) with a total of 3,682 patients met the inclusion criteria. The pooled mean difference (MD) suggested significantly elevated D-dimer levels in patients who died versus those survived (MD 6.13 mg/L, 95% CI 4.16 − 8.11, p <0.001). Similarly, the pooled mean D-dimer levels were significantly elevated in patients with severe COVID-19 infection (MD 0.54 mg/L, 95% CI 0.28 − 0.8, p< 0.001). In addition, the risk of mortality was four-fold higher in patients with positive D-dimer vs negative D-dimer (RR 4.11, 95% CI 2.48 − 6.84, p< 0.001) and the risk of developing the severe disease was two-fold higher in patients with positive D-dimer levels vs negative D-dimer (RR 2.04, 95% CI 1.34 − 3.11, p < 0.001).

**Conclusion:** Our meta-analysis demonstrates that patients with COVID-19 presenting with elevated D-dimer levels have an increased risk of severe disease and mortality.

## Introduction

The 2019 novel Coronavirus (2019-nCoV) or Severe Acute Respiratory Syndrome Corona Virus 2 (SARS-CoV-2), now declared a pandemic, first originated in December 2019 in Wuhan city of Hubei province, China and has since caused a significant impact on mankind ^1^. As of April 24th, 2020, 2.6 million individuals have been infected with SARS-CoV-2 in 213 countries worldwide, and 181,938 lives have been lost ^2^. On December 31st, 2019, China reported the outbreak to the World Health Organization (WHO). Subsequently, WHO officially declared the Coronavirus disease 2019 (COVID-19) epidemic as a public health emergency of international concern.^3^. The clinical features of COVID-19 vary from asymptomatic cases to severe infection, causing acute respiratory distress syndrome (ARDS), multisystem organ dysfunction, and death ^4^.

The overall case fatality rate (CFR) for COVID-19 was reported at about 2% in China. Still, it was noted to be higher at 7.2% in Italy, which was felt secondary to the higher mean age of the overall population ^5^. The CFR is significantly high in patients with severe COVID-19 infection with CFR as high as >50% in patients admitted to the Intensive Care Unit (ICU) ^6^. Due to high mortality in critically ill COVID-19 patients, the detection of biomarkers which may help identify them earlier in their course of illness can be crucial. D-dimer is one such biomarker that has emerged as an important prognostic tool, with elevated levels in critically ill patients and those deceased. In this systematic review, we aimed to investigate the prognostic role of admission D-dimer levels in patients hospitalized with COVID-19.

## Methods

### Search strategy

The reporting of this systematic review and meta-analysis complies with PRISMA (Preferred Reporting Items for Systematic Reviews and Meta-Analysis) guidelines (**Supplement Table 1**) **^7^**.

The initial search strategy was developed by two authors (SS and SP). We performed a systematic search, without language restriction, using PubMed, EMBASE, SCOPUS, Google Scholar, and two preprint servers (https://www.medrxiv.org/ and https://www.ssrn.com/index.cfm/en/coronavirus/) from inception to April 16th, 2020, for studies that reported D-dimer levels in COVID-19 patients. We utilized the “related articles” function in PubMed to find relevant articles that were missed by the initial search. In addition, reference lists of the included studies were hand-searched to further locate relevant articles that were missed in the primary search. We used the following keywords and medical subject heading: "COVID-19", "SARS-CoV-2", "Wuhan coronavirus", "Coronavirus 2019", "2019 n-CoV", "D-dimer", "laboratory".

### Study Selection and data extraction

To be included in our systematic review and meta-analysis the study had to fulfill the following criteria: (1) reported D-dimer levels in COVID-19 patients according to severity or include mortality as a clinical outcome; (2) included human subjects. (3) studies in English language. Single-arm studies, case reports, editorial, or systematic reviews were excluded. Two investigators (SS and SP) independently performed the literature search and screened all titles and full-text versions of all relevant studies that met study inclusion criteria.

The data from included studies were extracted using a standardized protocol and a data extraction form. Any discrepancies between the two investigators were resolved with a consultation with the senior investigator (JG). Two independent reviewers (SS and SP) extracted the following data from the eligible studies: author name, study design, publication year, follow-up duration, number of patients, age, gender, diabetes mellitus (DM), hypertension (HTN), coronary artery disease (CAD), acute cardiac injury, arrhythmias, shock, and outcomes. The Newcastle Ottawa Risk bias assessment tool was used to appraise the quality of the included studies (**Supplement Table 2**).

## Outcomes

### Clinical outcomes

The primary outcome of interest in our study was all-cause mortality and severity of COVID-19.

### Statistical Analysis

Mantel-Haenszel risk ratio (RR) random-effects model (DerSimonian and Laird method) was used to summarize data between the groups ^8^. The D-dimer levels in the studies were reported as median and Interquartile Range (IQR). We used the Wan method to estimate the mean and standard deviations ^9^. We then calculated the pooled difference in means (MD) to evaluate the association of levels of D-dimer between the groups. Higgins I-squared (*I^2^*) statistic was used to assess the test of heterogeneity. A value of *I^2^* of 0–25% represented insignificant heterogeneity, 26–50% represented low heterogeneity, 51–75% represented moderate heterogeneity, and more than 75% represented high heterogeneity ^10^. A pre-specified random-effects meta-regression analysis was conducted for the primary outcome in relation to the baseline demographics, comorbid condition, biomarkers to test the relationship between D-dimer and disease severity, and all-cause mortality. Publication bias was formally assessed using funnel plots and Egger’s linear regression test of funnel plot asymmetry. A two-tailed p < 0.05 was considered statistically significant. Statistical analysis was performed using Comprehensive Meta-Analysis version 3.0 (Biostat Solutions, Inc. [BSSI], Frederick, Maryland).

## Results

### Search results

A total of 920 citations were identified during the initial search (**Figure 1**). Nine hundred and two records were excluded. After a detailed evaluation of these studies, 12 studies met the inclusion criteria. We included six manuscripts from 2 pre-print servers (https://www.medrxiv.org/ and https://www.ssrn.com/index.cfm/en/coronavirus/), to accommodate the rapidly evolving nature of information for COVID. We acknowledge that the manuscripts from these two sources are not peer-reviewed. Eighteen articles including 3,682 patients were included in the final analysis.

**Figure 1:**
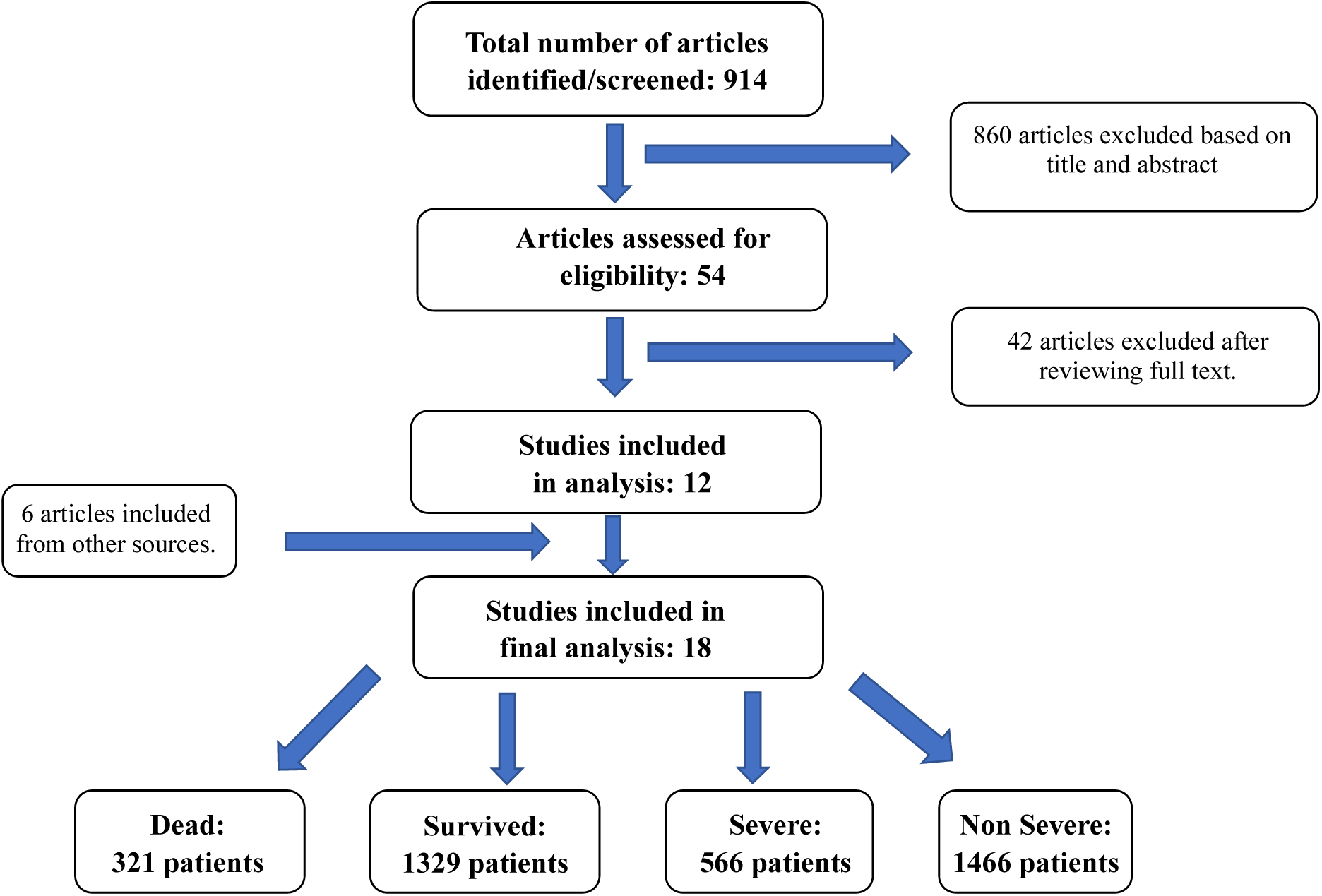
Flow Diagram illustrating the systematic search of studies

### Study characteristics

This systematic review and meta-analysis of 18 studies incorporated a total of 3,682 patients. Six articles compared D-dimer levels upon admission in dead versus survived patients ^11–16^, one article compared patients with elevated D-dimer level with normal D-dimer level ^17^ and 11 articles compared severe versus non-severe COVID-19 patients ^18–28^. All studies were retrospective ^12–27^ except two which were prospective ^11, 28^ and all were conducted in China, in the year 2020.

Positive d-dimer was defined as levels ≥0.5mg/L. Severe COVID-19 disease was defined as patients with respiratory rate ≥ 30 beats/minute (resting state) or mean oxygen saturation of ≤93% on room air or an arterial blood oxygen partial pressure (PaO2)/oxygen concentration (FiO2) ≤ 300 mm Hg and was consistent across all studies. The severe group included patients with severe COVID-19 and those needing ICU care for acute respiratory failure requiring mechanical ventilation, or for shock, or multiorgan failure. The acute cardiac injury was defined as an elevation in cardiac troponin or new changes of ischemia on electrocardiography (ECG) or new wall motion abnormalities on an echocardiogram.

**Table 1** summarizes the baseline characteristics of 6 studies which compared dead versus survived patients and 1 study which compared patients with elevated D-dimer vs normal D-dimer level. Among the 6 studies which compared dead versus survived patients, the mean age of the study population in this group was 62.5±14.8 years and 56.3% were males. Overall, hypertension (HTN) was the most common comorbidity (36.6%), followed by diabetes (DM) (16.8%) and coronary artery disease (CAD) (11.7%). The acute cardiac injury was present in 19.3% of patients while shock was observed in 8.9% of patients. New-onset arrhythmias of some form were observed in 12% of patients.

**Table 1:** Baseline characteristics of studies included in the meta-analysis comparing COVID-19 infected patients who died versus who survived

| Study Name | Study Type | Country | Study Period | Age (Y) | Male | Groups | N | Diabetes | Hypertension | Coronary Artery Disease | Acute cardiac injury | Arrythmias | Shock |
| --- | --- | --- | --- | --- | --- | --- | --- | --- | --- | --- | --- | --- | --- |
| Zhang, L. | Retrospective | China | Jan 12- March 15, 2020 | 62 (IQR 48-69) | 169 | Overall | 343 | 47 | 76 | 19 | NR | NR | NR |
| Wang, L. | Retrospective | China | Jan 1- Feb 6, 2020 | 69(IQR 65-76) | 166 | Survived | 274 | 43 | 106 | NR | 31 | 22 | 5 |
|  |  |  |  |  |  | Dead | 65 | 11 | 32 | NR | 39 | 13 | 3 |
| Tang, N. | Retrospective | China | Jan 1-Feb 13, 2020 | 65.1±12.0 | 268 | Survived | 315 | NR | NR | NR | NR | NR | NR |
|  |  |  |  |  |  | Dead | 134 | NR | NR | NR | NR | NR | NR |
| Zhou, F. | Retrospective | China | Dec 29, 2019 -Feb 1, 2020 | 56(IQR 46-67) | 119 | Survived | 137 | 19 | 32 | 2 | 1 | NR | NR |
|  |  |  |  |  |  | Dead | 54 | 17 | 26 | 13 | 32 | NR | 38 |
| Du, R. | Prospective | China | Dec 25, 2019-Feb 7, 2020 | 57.6 ± 13.7 | 97 | Survived | 158 | 27 | 45 | NR | NR | NR | NR |
|  |  |  |  |  |  | Dead | 21 | 6 | 13 | NR | NR | NR | NR |
| Cao, J. | Retrospective | China | Jan 3- Feb 1, 2020 | 54(IQR 37-67) | 53 | Survived | 85 | 5 | 17 | NR | 3 | 6 | 3 |
|  |  |  |  |  |  | Dead | 17 | 6 | 11 | NR | 12 | 12 | 7 |
| Zhang, F. | Retrospective | China | Dec 25, 2019-Feb 15, 2020 | 70.58±13.38 | 33 | Survived | 31 | 5 | 20 | 9 | NR | NR | NR |
|  |  |  |  |  |  | Dead | 17 | 5 | 12 | 4 | NR | NR | NR |

N = number; Y = years; IQR = interquartile range; NR = not reported

**Table 2** summarizes the baseline characteristics of 11 studies that compared severe versus non-severe COVID-19 patients. The mean age of the study population in this group was 49.9±17.2 years and 54.6% were males. Overall, HTN was the most common comorbidity (18.8%), followed by DM (9.2%) and CAD (3.9%). The acute cardiac injury was present in 11% of patients while shock was observed in 3.6% of patients, of which, 2% of patients have septic shock while it was undefined in other patients.

**Table 2:** Baseline characteristics of studies included in the meta-analysis comparing severe versus non-severe COVID-19 infected patients

| Study Name | Study Type | Country | Study Period | Age (Y) | Male | Groups | N | Diabetes | Hypertension | Coronary Artery Disease | Acute cardiac injury | Arrythmias | Shock |
| --- | --- | --- | --- | --- | --- | --- | --- | --- | --- | --- | --- | --- | --- |
| Deng, Q. | Retrospective | China | Jan 6- Feb 20, 2020 | 65(IQR 49-70.8) | 57 | Non-severe | 45 | 5 | 12 | 4 | 3 | NR | NR |
|  |  |  |  |  |  | Severe | 67 | 14 | 24 | 11 | 39 | NR | NR |
| Chen, G. | Retrospective | China | Dec-Jan 27, 2020 | 56(IQR 50-65) | 17 | Non-severe | 10 | 1 | 1 | NR | NR | NR | NR |
|  |  |  |  |  |  | Severe | 11 | 2 | 4 | NR | 1 | NR | 1 |
| Zhang, J. | Retrospective | China | Jan 16-Feb 3, 2020 | 57 (range 25-87) | 71 | Non-severe | 82 | 9 | 20 | 3 | NR | NR | NR |
|  |  |  |  |  |  | Severe | 58 | 8 | 22 | 4 | NR | NR | NR |
| Qian, G. | Retrospective | China | Jan 20-Feb 11, 2020 | 50(IQR 36.5-57) | 37 | Non-severe | 82 | 8 | 15 | NR | NR | NR | NR |
|  |  |  |  |  |  | Severe | 9 |  |  | NR | NR | NR | NR |
| Cai, Q. | Retrospective | China | Jan 11-Feb 09, 2020 | 47(IQR 33-61) | 149 | Non-severe | 240 | 19 | 38 | NR | 5 | NR | NR |
|  |  |  |  |  |  | Severe | 58 |  |  | NR | 15 | NR | NR |
| Ji, D. | Prospective | China | Jan 20-Feb 16, 2020 | 43.6±17.1 | 31 | Non-severe | 34 | NR | NR | NR | NR | NR | NR |
|  |  |  |  |  |  | Severe | 15 | NR | NR | NR | NR | NR | NR |
| Lu, H. | Retrospective | China | Jan 20- Feb 19, 2020 | NR | NR | Non-severe | 243 | 15 | 42 | 10 | NR | NR | NR |
|  |  |  |  |  |  | Severe | 22 | 6 | 10 | 4 | NR | NR | NR |
| Ma, K. | Retrospective | China | Jan 21-March 2, 2020 | 48(IQR 42.3-62.5) | 48 | Non-severe | 64 | 3 | 8 | NR | NR | NR | NR |
|  |  |  |  |  |  | Severe | 20 | 7 | 4 | NR | NR | NR | NR |
| Hu, L. | Retrospective | China | Jan 8- Feb 20, 2020 | 61(range 23-91) | 166 | Non-severe | 151 | 14 | 39 | NR | 2 | 18 | 4 |
|  |  |  |  |  |  | Severe | 172 | 33 | 66 | NR | 22 | 80 | 39 |
| Wan, S. | Retrospective | China | Jan 23- Feb 8, 2020 | 47(IQR 36-55) | 72 | Non-severe | 95 | 3 | 9 | NR | 8 | NR | 0 |
|  |  |  |  |  |  | Severe | 40 | 9 | 4 | NR | 2 | NR | 1 |
| Guan, W. | Retrospective | China | Dec 11, 2019- Jan 29, 2020 | 47(IQR 35-58) | 637 | Non-severe | 926 | 53 | 124 | 17 | NR | NR | 1 |
|  |  |  |  |  |  | Severe | 173 | 28 | 41 | 10 | NR | NR | 11 |

N = number; Y = years; IQR = interquartile range; NR = not reported

### All-cause mortality

The data for D-dimer levels were available in 5 studies ^11, 13–16^. The pooled mean D-dimer levels were significantly elevated in patients who died versus those who survived (MD 6.13 mg/L, 95% CI 4.16 − 8.11, p ≤ 0.001, *I^2^* = 81.41%) (**Figure 2**). No publication bias was observed (Egger’s P = 0.39, **Supplement Figure 1)**. Meta-regression analysis demonstrated no significant associations were found between age, male sex, hypertension, diabetes, coronary artery disease, C-Reactive Protein, and troponins in COVID-19 infected patients who died versus who survived (**Table 3**).

**Figure 2:**
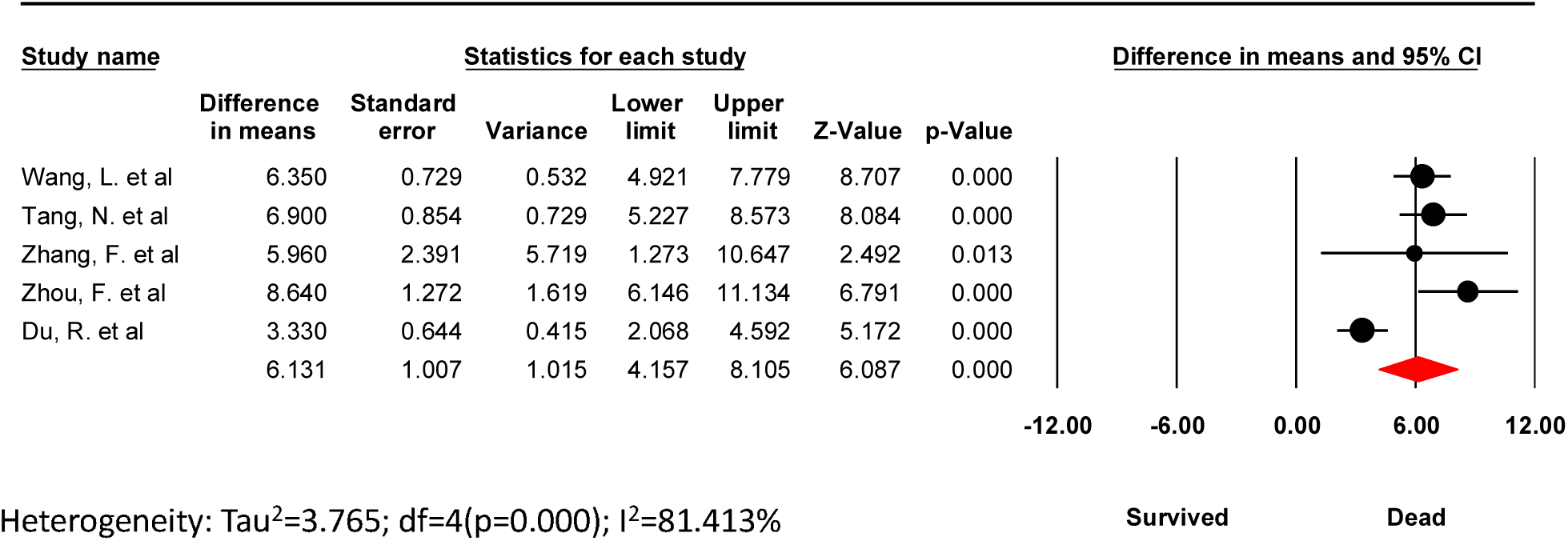
D-Dimer levels. The Forest plot for pooled difference in mean D-Dimer levels in dead versus survived COVID-19 patients.

**Table 3:** Meta regression of baseline characteristics with mean difference in D-dimer levels in COVID-19 patients – dead versus survived

| Dead vs survived COVID-19 |  | Meta regression |  |  |  |  |  |
| --- | --- | --- | --- | --- | --- | --- | --- |
|  | *Difference in Means*<br>*(95% CI)* | *Age* | *Male* | *Hypertension* | *Diabetes* | *CAD* | *Troponin* |
| D-Dimer levels | 6.13 (4.16 – 8.11),<br>P<0.001 | β: 0.02, p=0.91 | β: 0.05, p=0.62 | β: -0.002, p=0.98 | β: 0.15, p=0.82 | β: -0.12, p=0.25 | β: 201.41, p=0.53 |

CI- confidence interval, CAD- coronary artery disease

The risk of mortality was four-fold in patients with positive D-dimer vs. negative D-dimer (21% vs 4.9%, RR 4.11, 95% CI 2.48 − 6.84, p≤ 0.001, respectively). Test of heterogeneity was non-significant (*I*^2^= 0%) (**Figure 3**). No publication bias was observed (Egger’s P = 0.26, **Supplement Figure 2**).

**Figure 3:**
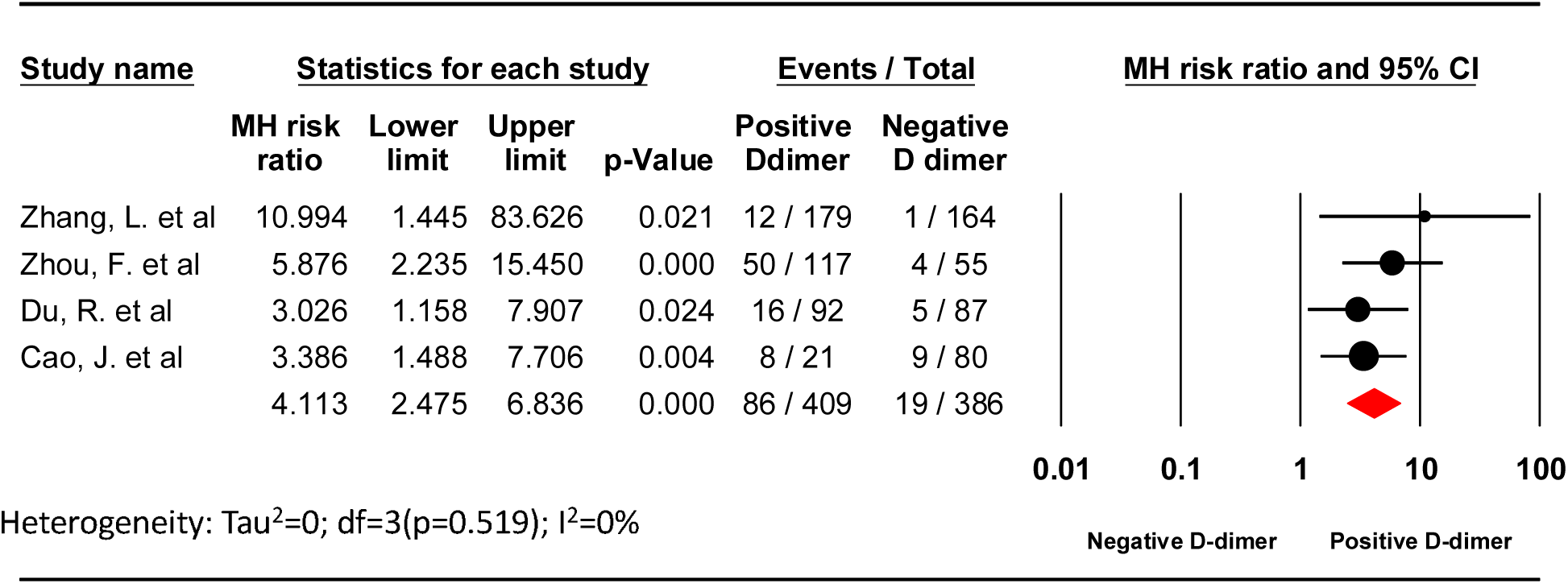
All-cause mortality. The Forest plot shows the outcomes of the individual trials as well as the aggregate.

### Severity of COVID-19

The data for D-dimer levels were available in 9 studies ^19–25, 27, 28^. The pooled mean D-dimer levels were significantly elevated in patients with severe COVID-19 infection (MD 0.54 mg/L, 95% CI 0.28 − 0.8, p≤ 0.001, *I^2^*= 90.74%) **(Figure 4A)**. No publication bias was observed (Egger’s P = 0.13, **Supplement Figure 3)**. Meta-regression analysis showed a significant association between CAD, C-Reactive Protein, and severe COVID-19 disease, but the results were not significant for age, male sex, comorbidities (hypertension, diabetes, troponin levels) (**Table 4**, **Figures 4B and 4C**).

**Figure 4:**
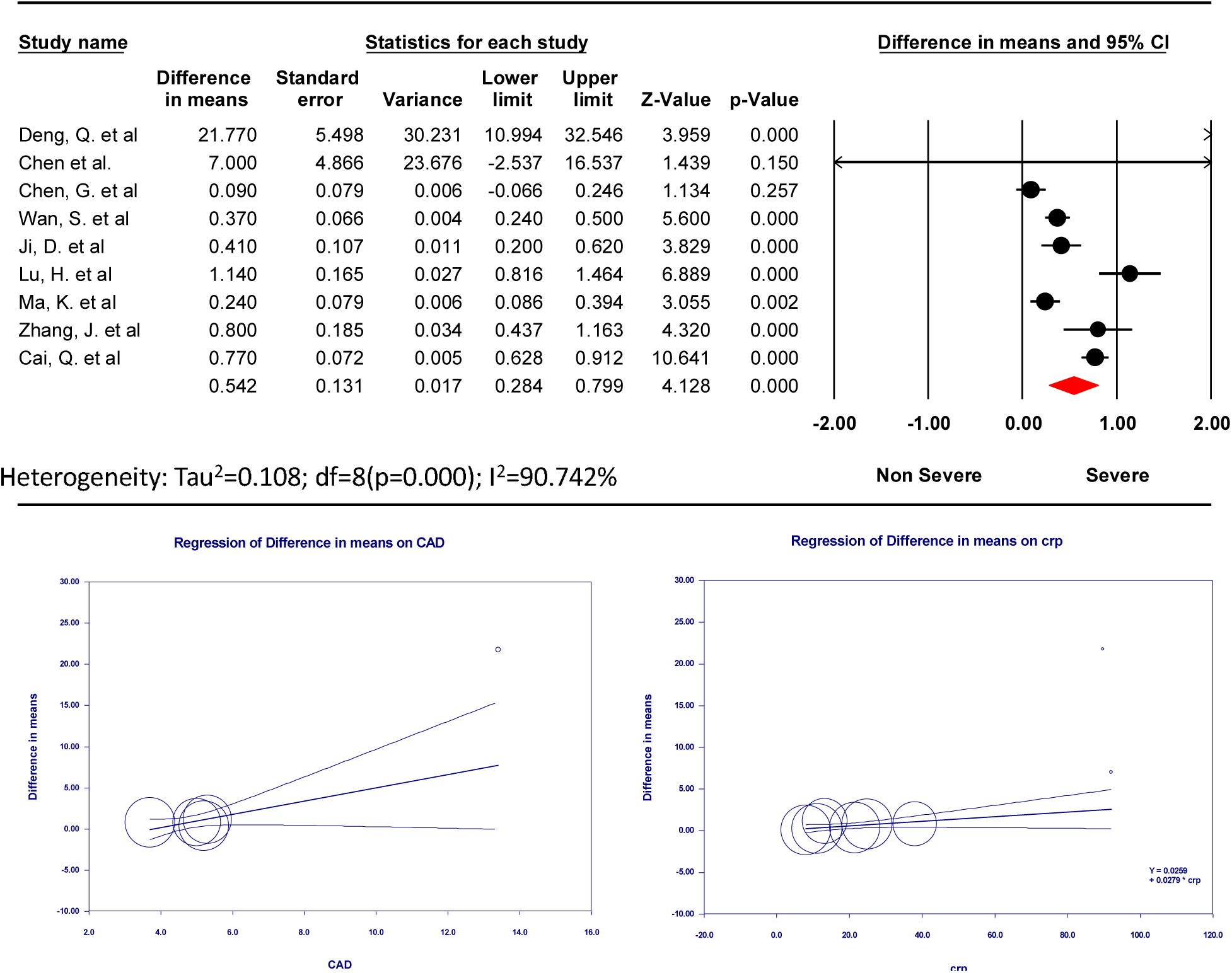
Disease severity. **(A)** The Forest plot for pooled difference in mean D-Dimer levels in severe versus non-severe COVID-19 patients, followed by random-effects meta-regression analysis plots depicting the relationship between mean differences in D-Dimer levels (on y-axis) and **(B)** Coronary Artery disease (CAD) and **(C)** C-Reactive Protein (CRP). Each included study is represented by a circle, the size of which is proportional to its respective weight in the analysis. The line indicates the predicted effects (regression line). There was significant association between CAD (β = 0.8, P = 0.02), and CRP levels (β = 0.02, P = 0.03) and mean differences in D-Dimer levels.

**Table 4.** Meta regression of baseline characteristics with mean difference in D-dimer levels in severe versus non-severe COVID-19 infected patients

| Severe vs non-severe COVID-19 |  | Meta regression |  |  |  |  |  |  |
| --- | --- | --- | --- | --- | --- | --- | --- | --- |
|  | *Difference in Means*<br>*(95% CI)* | *Age* | *Male* | *Hypertension* | *Diabetes* | *CAD\** | *CRP\** | *Troponin* |
| D-Dimer levels | 0.54 (0.28 – 0.80),<br>P<0.001 | β: 0.03, p=0.31 | β: 0.008, p=0.68 | β: 0.03, p=0.2 | β: -0.01, p=0.84 | β: 0.8, p=0.02 | β: 0.02, p=0.03 | β: 105.63, p=0.06 |

CI- confidence interval, CAD- coronary artery disease, CRP- C reactive protein, \* indicates statistically significant value.

The risk of developing the severe disease was two-fold higher in patients with positive D-dimer levels vs negative D-dimer (40.74% vs. 21.98%, RR 2.04, 95% CI 1.34 − 3.11, p ≤ 0.001, *I^2^*=81.83%, respectively) (**Figure 5**). No publication bias was observed (Egger’s P = 0.16, **Supplement Figure 4**). A sensitivity analysis was performed by removing one study at a time (n-1 analysis) to investigate the significant heterogeneity. No significant change in the findings was observed with the sensitivity analysis.

**Figure 5:**
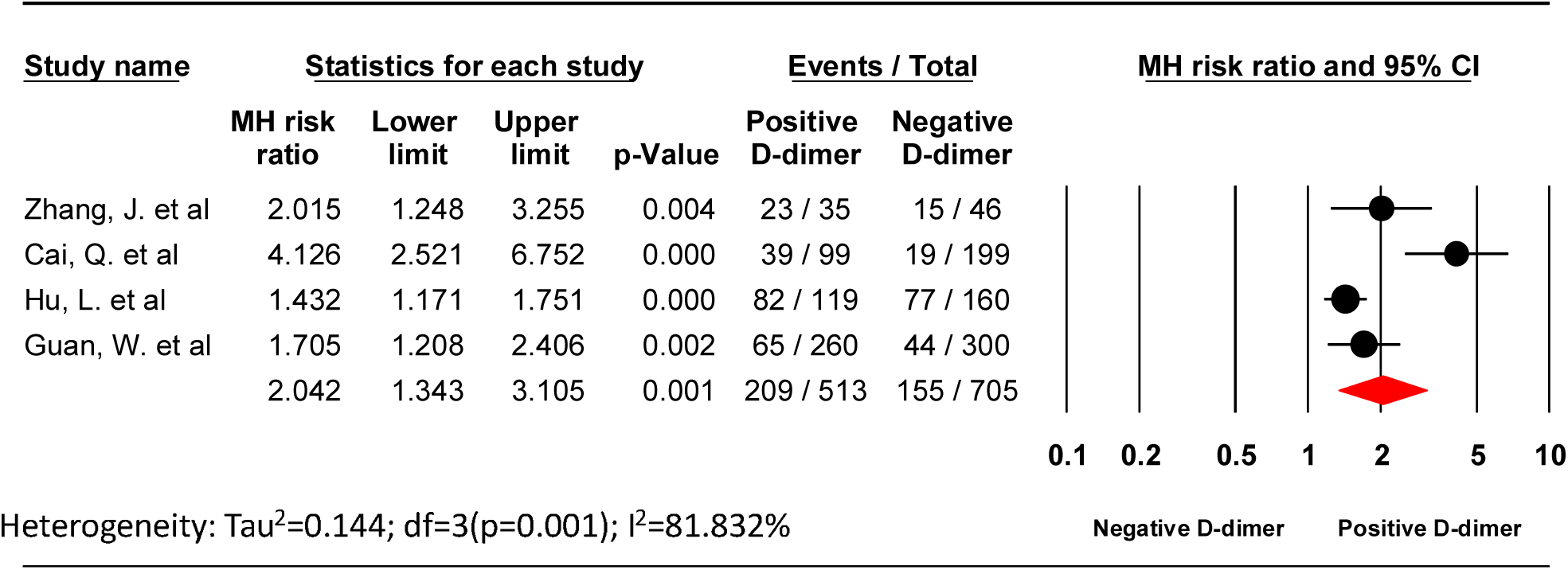
The Forest plot demonstrating the risk ratio of positive D-Dimer with severity.

## Discussion

Elevated D-dimer is one of the abnormal laboratory parameters in patients with COVID-19 infection. D-dimer is the fibrin degradation products released upon cleavage of crosslinked fibrin by plasmin ^29^. Historically, the role of D-dimer is limited due to its nonspecificity, with elevated levels are often seen with advanced age, African American race, female sex, active malignancy, surgery, pregnancy, immobility, cocaine use, connective tissue disorders, end-stage renal disease and prior thromboembolic disease ^30^. The D-dimer is routinely utilized clinically in diagnosing disseminated intravascular coagulation (DIC) and those with low pretest probability for deep vein thrombosis (DVT) and pulmonary embolism (PE) ^29^.

More recently, D-dimer has been explored to identify patients thought to develop severe COVID-19 infection earlier in their course of illness. Elevated D-dimer level was discovered in about 36%-47% of hospitalized patients with COVID-19 infection, the majority of whom are those with severe COVID-19 infection ^31^. A prior meta-analysis comprising of 4 studies showed a higher D-dimer level in patients with severe COVID-19 infection compared to those with the non-severe disease ^32^. However, this meta-analysis was limited by relatively smaller sample size. Also, it did not answer an important question regarding the prognostic value of D-dimer in predicting severe COVID-19 infection and mortality. Our meta-analysis comprising of 18 studies evaluated the prognostic role of D-dimer in COVID-19 and is the largest to date to the best of our knowledge. The key findings of our pooled analysis are: 1) the D-dimer levels were higher in patients with severe COVID-19 infection and those who succumbed to death compared to non-severe disease and those who survived, respectively after adjusting for age, comorbid condition, CRP levels; 2) patients with elevated D-dimer levels were at increased risk of developing severe COVID-19 infection and increased all-cause mortality compared to those with normal D-dimer levels.

Zhou et al. ^13^ reported that D-dimer level >1 mg/L on admission was independently associated with increased odds of mortality, findings that echoes in our pooled analysis as well. Also, patients with advanced age, higher Sequential Organ Failure Assessment (SOFA) score, elevated troponin, and B-type natriuretic peptide (BNP) have been associated with poor outcomes and mortality in COVID-19 infection ^13, 33, 34^. Furthermore, using a higher cutoff value of D-dimer (levels > 2mg/L) predicted in-hospital mortality even better as noted by Zhang at el. ^17^ with a sensitivity of 92.3% and a specificity of 83.3% after adjusting for age, gender and comorbidities. Besides, studies have shown that rising D-dimer levels during the course of hospitalization are associated with worst long term outcomes ^12, 13^. Additionally, COVID-19 patients with one or more comorbidities [HTN, DM, and cardiovascular diseases] are associated with adverse outcomes (i.e. severe COVID-19 disease and/or mortality) ^35–37^. In our pooled analysis, patients with severe COVD-19 infection had significantly elevated D-dimer levels, with an increasing prevalence of HTN, DM, and CAD.

There has been upcoming evidence regarding an increased incidence of venous thromboembolic events (VTE), including DVT and PE in patients with severe COVID-19 infection ^38^. Besides, disseminated intravascular coagulation (DIC) has been increasingly reported in these patients. Tang et al. found a 3.5-fold increase in D-dimer levels in those who died and 71% of them met the International Society on Thrombosis and Hemostasis (ISTH) criteria for DIC compared to 0.6% only among those who survived ^39^. Similarly, another study proposed that D-dimer level >1.5mg/L may help detect VTE events with a sensitivity of 85.0% and specificity of 88.5%, however, results should be interpreted with caution due to small sample size and lack of external validation ^40^. It is unclear at this time if this is a direct consequence of SARS-CoV-2 infection or a due to cytokine storm resulting in systemic inflammatory response syndrome (SIRS), as seen in other viral infections ^41–44^. A similar pattern of changes in coagulation cascade with increased prothrombotic state and incidences of DVT and PE were also noted with coronavirus responsible for Middle Eastern Respiratory Syndrome (MERS-CoV) and SARS-CoV-1 ^45^. The risk of VTE is generally high in critically ill patients, but the risk appears to be higher in patients infected with SARS-CoV-2.

Due to several reasons for D-dimer elevation in these patients, imaging studies to diagnose DVT or PE should only be pursued if clinically warranted ^40^. High clinical suspicion for DVT or PE is warranted in patients with elevated D-Dimer (more so in > 2mg/dl), as failure to treat may result in adverse clinical outcomes ^17^. Thus, it is possible that patients who remained untreated for this catastrophe condition, accounted for adverse clinical outcomes as noted in our pooled analysis. However, no such information was available from the studies included in our analysis to evaluate for this difference. Also, performing imaging like CT angiogram of the chest can often be challenging in these patients due to isolation precautions and unstable hemodynamics, inability to administer intravenous contrast due to acute kidney injury and proning. In such cases, identification of new right ventricular dysfunction and/or enlargement on transthoracic echocardiogram can be useful. Perhaps, empirically treating all COVID-19 patients with intermediate or full (therapeutic) doses of anticoagulation to prevent microvascular thrombosis ^14, 46^ might be beneficial (provided a thorough risk-benefit assessment given these patients are also at risk of spontaneous bleeding) (however, our study was not designed to assess this difference). Besides that, it remains unclear at this time regarding the optimal dosing and duration in this patient population and hence needs to be explored further. Although, extended DVT prophylaxis with oral anticoagulation at discharge (for up to 45 days) may be reasonable in patients at higher risk for the thromboembolic event (i.e. active malignancy, immobility and elevated D-dimer level > two times the upper limits of normal) and lower bleeding risk ^47, 48^. Thus, using D-dimer levels as a surrogate marker for disease severity and underlying thromboembolic disease, especially, in COVID-19 patients who cannot get dedicated imaging might be beneficial.

Our study has a few important limitations. First, all studies included in our meta-analysis were from China, while the United States and Europe have the majority of COVID-19 cases currently. However, the preliminary reports from the United States and Europe have shown similar trends in COVID-19 infection in terms of clinical presentation and outcomes ^5, 49^. Our pooled analysis provides the best available data regarding trends of D-dimer levels in patients with COVID-19 infection and the likelihood of developing severe infection or mortality in patients with elevated D-dimer levels. Secondly, all studies included in our analysis were either prospective or retrospective reports, which is currently the best available evidence; and, therefore, subject to potential confounding and publication bias. Third, significant heterogeneity was observed between studies in our pooled analysis. Fourth, details on anticoagulation or trends of D-dimer over the course of hospitalization were not available. Finally, patient-level data to perform additional. detailed analyses are not available.

## Conclusion

Our meta-analysis demonstrates that patients with COVID-19 presenting with elevated D-dimer levels have an increased risk of severe disease and mortality.

## Data Availability

not applicable

---

**Supplement Table 1:** PRISMA checklist

**Supplement Table 2:** Risk of bias assessment of studies included in our meta-analysis using Newcastle-Ottawa Scale

**Supplement Figure 1: D-Dimer levels.** The funnel plot of studies included in the analysis assessing D-dimer levels in dead versus survived COVID-19 patients

**Supplement Figure 2: All-cause mortality.** The funnel plot of studies assessing the association of elevated D-dimer and all-cause mortality in COVID-19 patients.

**Supplement Figure 3:** The funnel plot of studies included in the analysis assessing D-dimer levels in severe versus non-severe COVD-19 infected patients.

**Supplement Figure 4:** The funnel plot of studies assessing the association of elevated D-dimer and COVID-19 disease severity.

